# Facility-based HIV self-testing strategies may substantially increase number of men and youth tested for HIV in Malawi: results from a data-driven individual-based model

**DOI:** 10.1101/2021.08.25.21262593

**Authors:** Brooke E Nichols, Alexandra de Nooy, Mariet Benade, Kelvin Balakasi, Misheck Mphande, Gabriella Rao, Cassidy W Claassen, Shaukat Khan, Christian Stillson, Naoko Doi, Kathryn Dovel

**Affiliations:** Department of Global Health, School of Public Health, Boston University, Boston, MA, USA; Health Economics and Epidemiology Research Office, Department of Internal Medicine, School of Clinical Medicine, Faculty of Health Sciences, University of the Witwatersrand, Johannesburg, South Africa; Department of Medical Microbiology, Amsterdam University Medical Centre, Amsterdam, the Netherlands; Right to Care, Johannesburg, South Africa; Partners in Hope, Lilongwe, Malawi; Tufts University, Boston, MA; Center for International Health, Education, and Biosecurity, University of Maryland School of Medicine, Baltimore, MD, USA; Division of Infectious Diseases, Department of Medicine, University of Zambia School of Medicine, Lusaka, Zambia; Clinton Health Access Initiative, Boston, MA, USA; Division of Infectious Diseases, Department of Medicine, University of California Los Angeles David Geffen School of Medicine, University of California Los Angeles

## Abstract

**Background:** Malawi is rapidly closing the gap in achieving the UNAIDS 95-95-95 targets, with 90% of people living with HIV in Malawi aware of their status. As we approach epidemic control, interventions to improve coverage will become more costly. There is therefore an urgent need to identify innovative and low-cost strategies to maintain and increase testing coverage without diverting resources from other HIV services.

**Methods:** A data-driven individual-based model was parameterized with data from a community-representative survey (sociodemographic, health service utilization, HIV testing history) of men and youth in Malawi (data collected 08/2019). 79 different strategies for the implementation of HIV self-testing (HIVST) and provider-initiated-testing-and-counselling at the outpatient department (OPD) were evaluated. Outcomes included percent of men/youth tested for HIV in a 12-month period, cost-effectiveness, and human resource requirements. Testing yield was assumed to be constant across the scenarios.

**Findings:** Facility-based HIVST offered year-round resulted in the greatest increase in proportion of men and youth tested in the OPD (from 45% to 72%-83%), was considered cost-saving for HIVST kit priced at $1.00, and generally reduced required personnel as compared to the status quo. At higher HIVST kit prices, and more relaxed eligibility criteria, all scenarios that considered year-round HIVST in the OPD remained on the cost-effectiveness frontier.

**Interpretation:** Facility-based HIVST is a cost-effective strategy to increase the proportion of men/youth tested for HIV and decreases the human resource requirements for HIV testing in the OPD-providing additional health care worker time for other priority health care activities.

**Funding:** FCDO; USAID

## INTRODUCTION

Among people living with HIV in Malawi, 90% were aware of their status, 87% of those who knew their status were on treatment, and 92% of those on treatment had achieved viral suppression – rapidly closing the gap in achieving the UNAIDS *95-95-95 by 2025* targets.^1^ Yet these successes do not translate to all populations – there are substantial disparities in the treatment cascade for sub-populations such as men and youth. Innovative strategies are needed to address these disparities, however, effective interventions become more costly as we approach epidemic control.^2^ There is an urgent need to identify innovative and low-cost strategies to maintain and increase testing coverage to close the final testing gap for men and youth without diverting resources from other HIV services.

Men and youth in Malawi are most often missed by routine HIV testing. Men are likely to be missed because their frequent entry point into health facilities – outpatient departments (OPD) – rarely results in routine provider-initiated-testing-and-counseling (PITC), unlike family planning or antenatal entry points frequented by women.^3^ One novel strategy to improve testing coverage in OPD is HIV self-testing (HIVST), whereby individuals conduct and interpret an HIV test on their own. Clients who receive a reactive HIVST result and disclose their result are then referred for professional-use testing following the national testing algorithm. HIVST is a promising strategy for scale-up in Malawi and across similar settings due to its simple procedure, low staffing requirements, high uptake and acceptability among men and youth, and low risk for adverse events.^4^ Among outpatients in Malawi, facility-based HIVST within private waiting spaces has proven to increase testing coverage compared to PITC.^5^ Facility-based HIVST may also be less costly and more effective than community-based self-testing strategies, as both men and youth do attend health facilities, and more specifically, OPD.^3,6^ Thus, implementation of a combination of HIVST and PITC within OPD has the potential to vastly increase testing coverage in Malawi and other similar settings, without incurring substantial additional cost.

Only a handful of studies have evaluated the impact of facility-based HIVST, and most are implemented over a brief time-period.^7,8^ It is possible that there is a reduction in the effectiveness of a facility-based HIVST strategy as the population to be tested saturates. Additionally, the above trials did not assess the proportion of community members likely to benefit from a facility-based HIVST strategy, leaving a critical gap in understanding the potential reach of facility HIVST at the population level. Fortunately, our recent community-representative survey of men and youth in Malawi has demonstrated that, while the proportion of men and youth who have tested for HIV in the past year is relatively low (45%), the majority of those surveyed had visited a health facility in the past year (82%) and attended as either a client (61% of visits) or as a guardian (meaning they attended to support the health care of others) (39% of visits).^3^ Most attended OPD (84% of visits).^3^ This indicates that a facility-based testing strategy has potential to improve community-level testing coverage among men and youth, since most individuals regularly attend facilities.

Through the creation of a data-driven individual-based model, parameterized with survey data of a community-representative survey of men and youth in Malawi, we modelled the impact of using facility-based HIV testing modalities to determine the most cost-effective strategy to increase the proportion of men and youth testing for HIV.

## METHODS

### Study population

This individual-based simulation model was parameterized using the results of a community-representative survey of Malawian 1,180 men (ages 15-64 years) and 300 young women (ages 15-24 years). The survey was conducted from August-October 2019 across 36 randomly selected villages in Central and Southern Malawi using a multi-staged sampling design. The parent study was designed to assess utilization of facility-based health services over the past two years, offer and uptake of HIV testing services, reasons for testing or not, and willingness to use an HIV self-test. Additional details regarding the parent study have been published elsewhere.^3^

Eligibility criteria included: (1) aged 15-64 years for men or aged 15-24 for women; (2) current resident of the participating village; and (3) spent >15 nights within the village in the past 30 days. Those self-reported as ever testing HIV-positive were excluded from the study since they fall outside the target population for HIV testing strategies. Random selection was stratified by village (n∼45 per village, although some villages had fewer than 45 men due to small village size) and age categories: young men (15-24 years, n=300); middle-aged men (25-39 years, n=425); older men (40+ years, n=425); and young women (15-24 years, n=300).

### Model development

Characteristics were assigned to each individual in the model based on survey results (n=1,480). Characteristic variables included age, sex, and numerous questions regarding facility attendance history, such as date and reason for recent facility visits, whether HIV testing was offered and/or accepted, and whether the visit was as a client or a guardian. The simulations were scaled by a factor of 100 to aid ease of interpretability for a total simulated population of 148,000.

### Facility attendance history

Survey data included date of facility attendance for the four most recent visits within the past 4-years (whether as client or guardian), as well as the total number of visits over a 24-month period prior to completing the survey. For this analysis, we only include visits within the past 12-months given likely recall bias for visits >12-months ago. For every visit date assigned, the visit reason, and whether the visit was as a client or a guardian, was recorded and assigned to the individual in the model. The individual simulations were programmed in MATLAB v9.7 (Natick, MA).

### Human resource calculations

We estimated the number of healthcare worker hours required to implement each facility-based testing scenario (time required per test reported in **Table 1**). Calculations were based on the staff time required to complete all tests within a given month, assuming an equal distribution of tests across all weekdays. Using different algorithms for PITC and HIVST, the total person time required to complete tests was determined, assuming six hours of patient-provider interaction per day per staff member and 21.5 working days per month, into a daily staff requirement for each scenario. The number of healthcare workers required in the month of peak testing was then calculated to ensure that the described staff requirements would be sufficient for all months under consideration. The algorithm for PITC and HIVST staff time are described below.

**Table 1.**
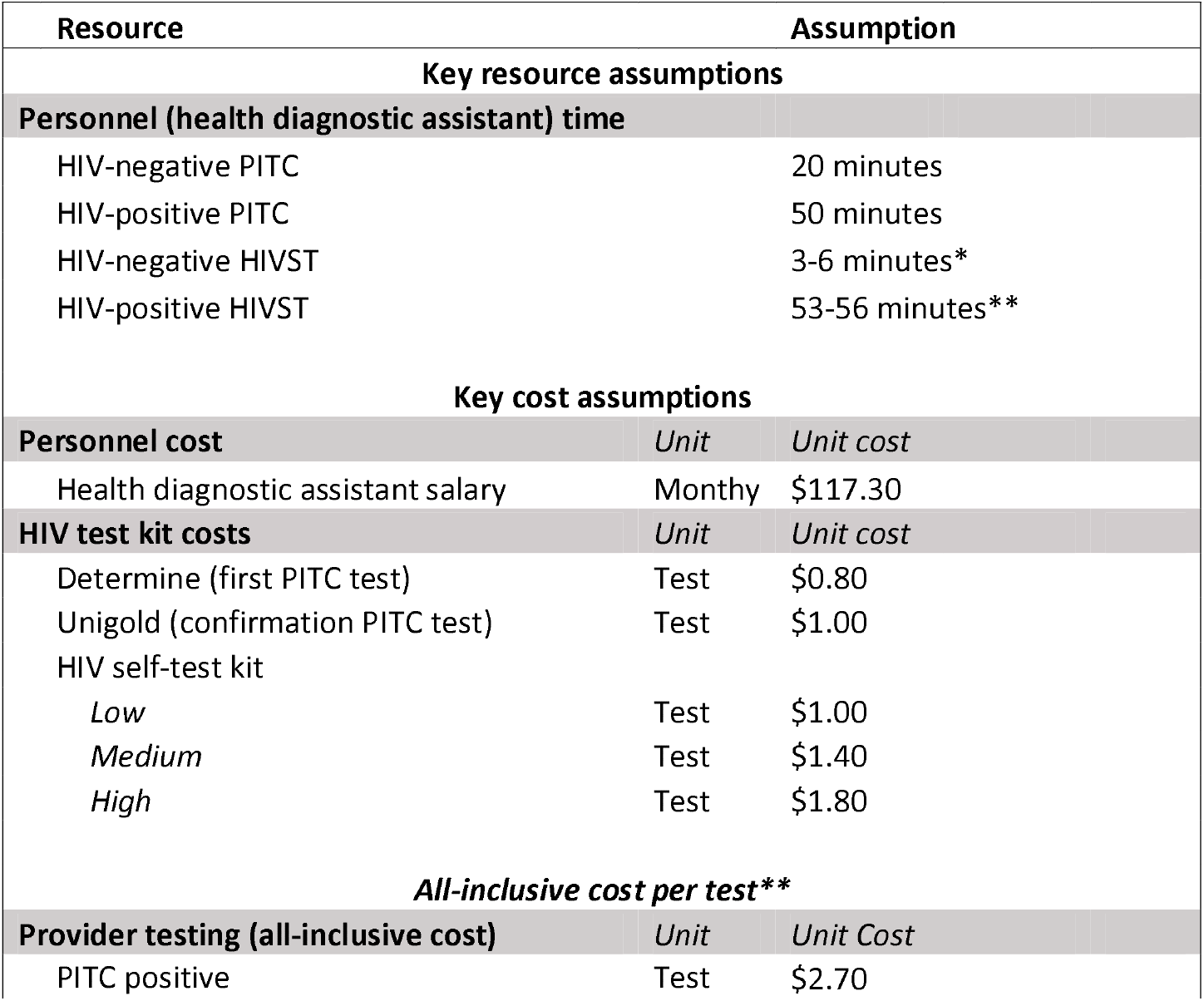
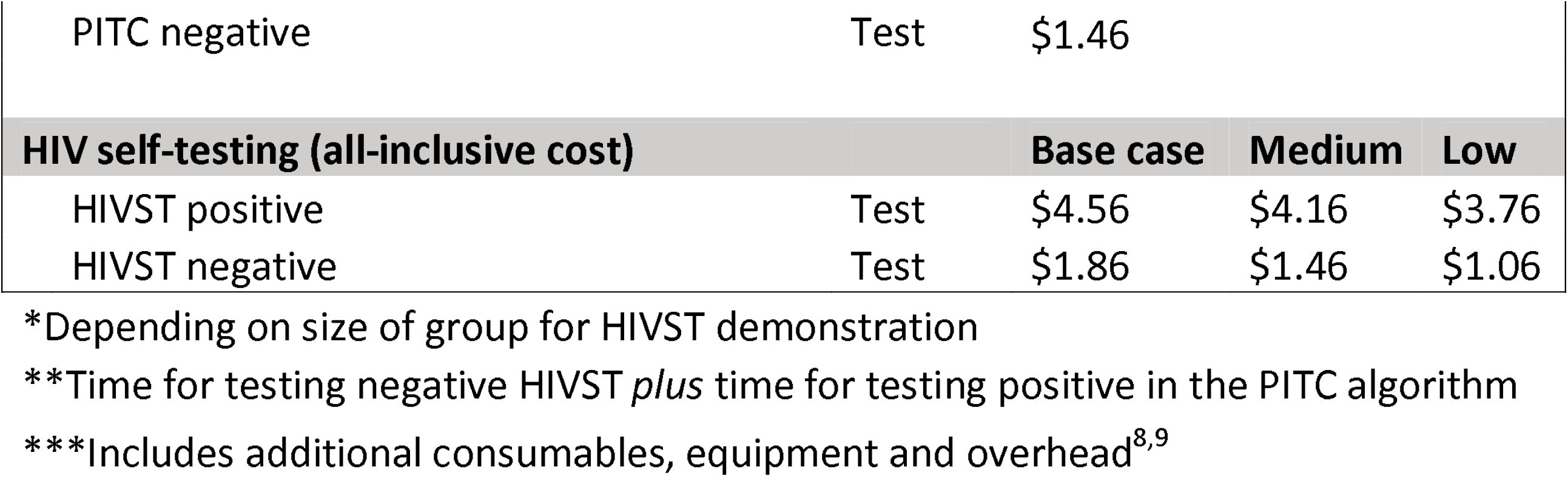
Key HIV testing cost and resource assumptions^8,9^

#### PITC Staff Time

Each negative test was assumed to require 20-minutes of staff interaction time, and each HIV-positive test to require 50-minutes of staff interaction time.^9^ Overall PITC staff time was taken as the summation of person-time for the number of positive and negative tests of the total cohort assigned a PITC test across the 12-month period. From the community representative survey, 15% of individuals refused PITC HIV testing when offered. This was also assigned in the individual simulations, and we assumed that when one of these individuals was assigned to PITC, that PITC would not be successful in testing the individual.^3^

#### HIVST Staff Time

Time calculations for self-testing assumed an implementation through group information sessions, with one staff member hosting each session, followed by self-test kit distribution. Similar to a recent cluster randomized trial of facility-based HIVST in Malawi, sessions were assumed to consist of a 30-minute HIVST demonstration, 10-minutes for questions and test distribution, and an additional 2-minutes for every individual in the group for test kit distribution.^8^ We ranged the number of HIVST demonstrations and question sessions per day between 2-6, with their person-time divided by the number of people tested on average during a day in that respective month. The test kit distribution time was multiplied by the average number of people testing across the month. Those who tested positive with HIVST were then assumed to enter the PITC testing algorithm from the beginning and therefore an additional 50-minutes of testing person-time required. In scenarios in which HIVST was assigned, 86% of adult men, 88% of young men, and 88% of young women were assumed to accept an HIVST based on the community survey. We assumed that HIVST refusers were offered a test through PITC, and that of those refusing HIVST, 96% accepted PITC.^3^

### Testing scenarios

We considered a set of scenarios to review the impact of different facility-based testing interventions. The scenarios included variations of the following:

1. Varying coverage of standard (professional use) PITC and HIVST from 0%-100%
2. Targeted to OPD clients, guardians, or both clients and guardians
3. Targeted to different time frames (highest OPD volume month, two months, or three months out of the year; or full 12 months)

A set of 79 mutually exclusive intervention combinations of the above three factors were considered (Table 2). Assignment of tests at each coverage level was performed using random number generation. For example, a coverage of 30% of PITC would be the percentage of all visits, at random, that were offered PITC. The final number of people tested is the number offered tested multiplied by the uptake of testing at the individual level, parameterized from the community survey. Scenarios where just three months were targeted for intervention were evaluated to test whether a more parsimonious testing algorithm could result in a similar number of people diagnosed. In Malawi, there is typically a three month period each year with an increase in OPD visits due to influenza infections or malaria, and these HIV testing scenarios reflect implementation during this part of the year where there are more OPD visits. For the full facility-based HIVST scenarios, where HIVST is offered all twelve months of the year, we also assessed different screening criteria for being eligible to test (defined as 12+ months since last test, 6+ months since last test, 3+ months since last test, and no criteria).

**Table 2.** Summary of combinations of facility-based HIV self-testing and provider-initiated testing and counseling scenarios tested.

| Testing type |  | Provider-initiated testing and counseling (PITC) |  |  |  |  |  | HIV self-testing (HIVST) |  |  |  |
| --- | --- | --- | --- | --- | --- | --- | --- | --- | --- | --- | --- |
| Number of months implemented |  | 12 months |  | 9 months |  | 3 months |  | 12 months |  | 3 months |  |
|  |  | Person Tested | Percentage* | Person Tested | Percentage* | Person Tested | Percentage* | Person Tested | Percentage* | Person Tested | Percentage* |
| Scenario | 1 | Client | 5-90% |  |  |  |  |  |  |  |  |
|  | 2 | Both | 5-90% |  |  |  |  |  |  |  |  |
|  | 3 |  |  | Both | 10% | Both | 15-90% |  |  |  |  |
|  | 4 |  |  | Both | 5-90% | Both | HIVST refusers** |  |  | Both | 100 |
|  | 5 | Client | 5-90% |  |  |  |  |  |  | Guardian | 100 |
|  | 6 |  |  | Both | 5-90% | Client | 5-90% |  |  | Guardian | 100 |
|  | 7 |  |  | Both | 5-90% | Guardian | 5-90% |  |  | Client | 100 |
|  | 8 | HIVST refusers** |  |  |  |  |  | Both | 100% |  |  |

\*Percentage of all visits covered with a test
\*\*PITC only offered to those who refused HIVST

Within the model design, there are noted elements which were randomly implemented. For each scenario, this related to when tests were assigned to visits. As such, to achieve a better overview of what typical implementation may look like, each scenario was iterated multiple times. In total, 100 iterations of each scenario were implemented and the average results for all iterations were exported for analysis.

### Scenario costing and cost-effectiveness analysis

To calculate the total costs for each scenario we accounted for the full cost of all test kits, including professional-use tests prescribed by Malawi’s HIV testing algorithm, Determine HIV 1/2 (Alere), Uni-Gold HIV (Trinity Biotech), and OraQuick HIVST (OraSure), staff time, consumables and overhead estimated from previous work.^8,9^ The unit costs for positive and negative tests by modality are presented in Table 1. To determine cost-effectiveness, we calculated the incremental cost-effectiveness ratio of each scenario at different HIVST kit prices-given that the kit price can affect the order of scenarios on the cost-effectiveness frontier. To enable comparability to the literature, we define effectiveness as individuals newly diagnosed with HIV.^9,10^ We have set a constant 2.5% testing yield across scenarios to enable the use of this outcome measure.^8^ We considered three different price-points of the HIVST kit to reflect the shifting market: $1.80 (base case), $1.40, and $1.00 per test kit. Testing scenarios that are on the cost-effectiveness frontier are reported separately. Costs are reported in 2018 USD collected as part of a previous facility-based HIVST trial in Malawi.^8,9^

### Ethics

The National Health Sciences Review Committee of Malawi (number 2338) and the University of California Los Angeles Institutional Review Board (number 20–001606) approved study activities.

## RESULTS

Of the 79 scenarios tested, 61 increased the proportion of men and youth tested in the past 12 months compared to the baseline PITC scenario (in which PITC is offered at approximately 50% of visits) (Table 3). The scenario that had the greatest increase in the proportion of men and youth tested was Scenario 8, offering facility-based HIVST all 12-months to both clients and guardians (and offering PITC to those who refuse HIVST, regardless of time since last HIV test), resulting in as much as a 79% increase in those tested within the past 12-months, from 46% baseline to 83% testing coverage. The next most effective scenario (Scenario 4) was PITC for clients and guardians at high coverage levels (>40% coverage) for 9 months of the year combined with three high OPD volume months of facility-based HIVST for men and youth clients and their guardians – resulting in up to a 77% increase in proportion tested (46% baseline to 81% tested within 12 months). High levels of PITC coverage (>40%) for both clients and guardians year round (Scenario 2) can result in a similar proportion of men and youth tested—as compared to a facility-based HIVST scenario (Scenario 8), however, PITC-only scenarios with high coverage also require large increases in personnel cost to administer additional tests (up to a 174% increase).

**Table 3.**
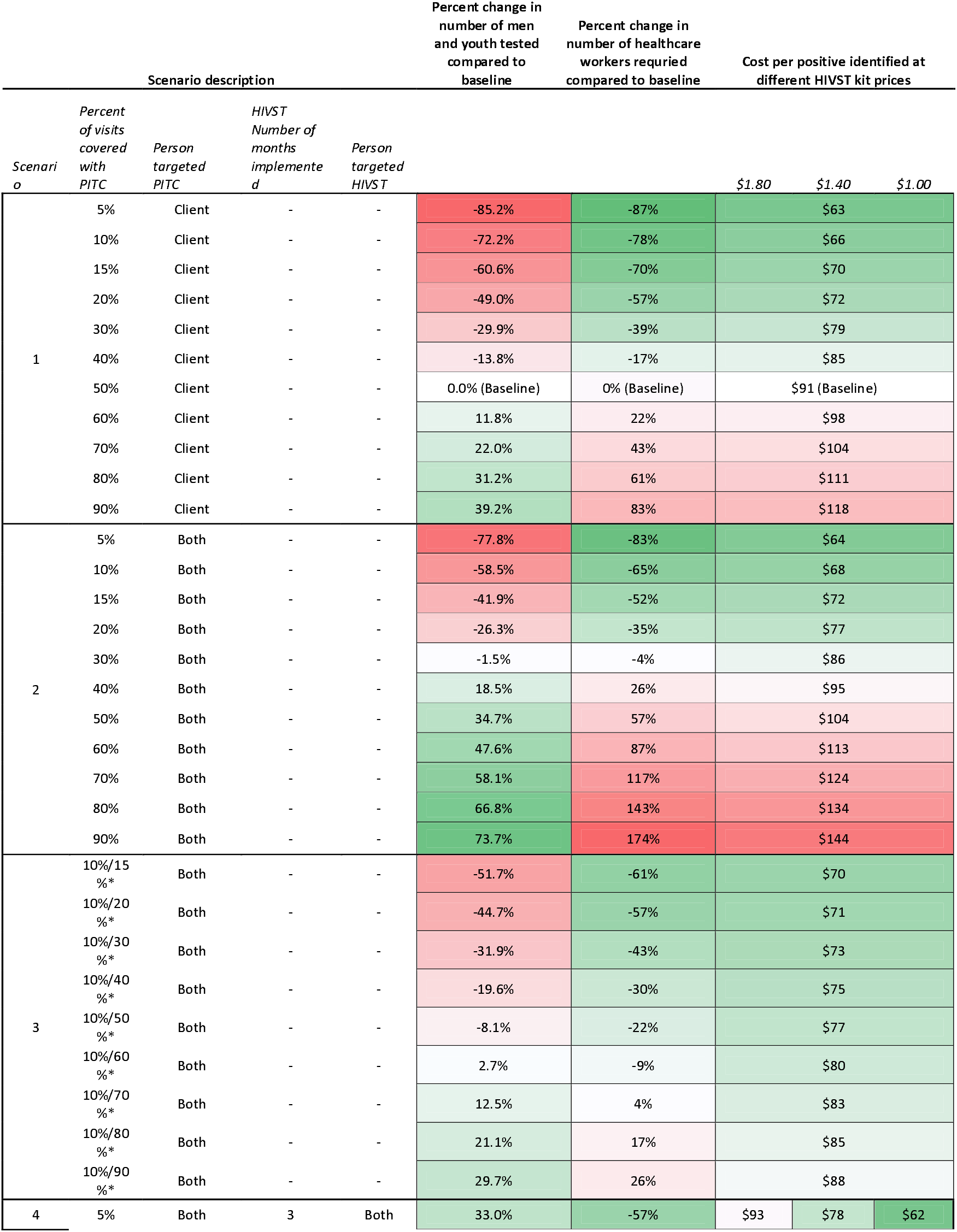
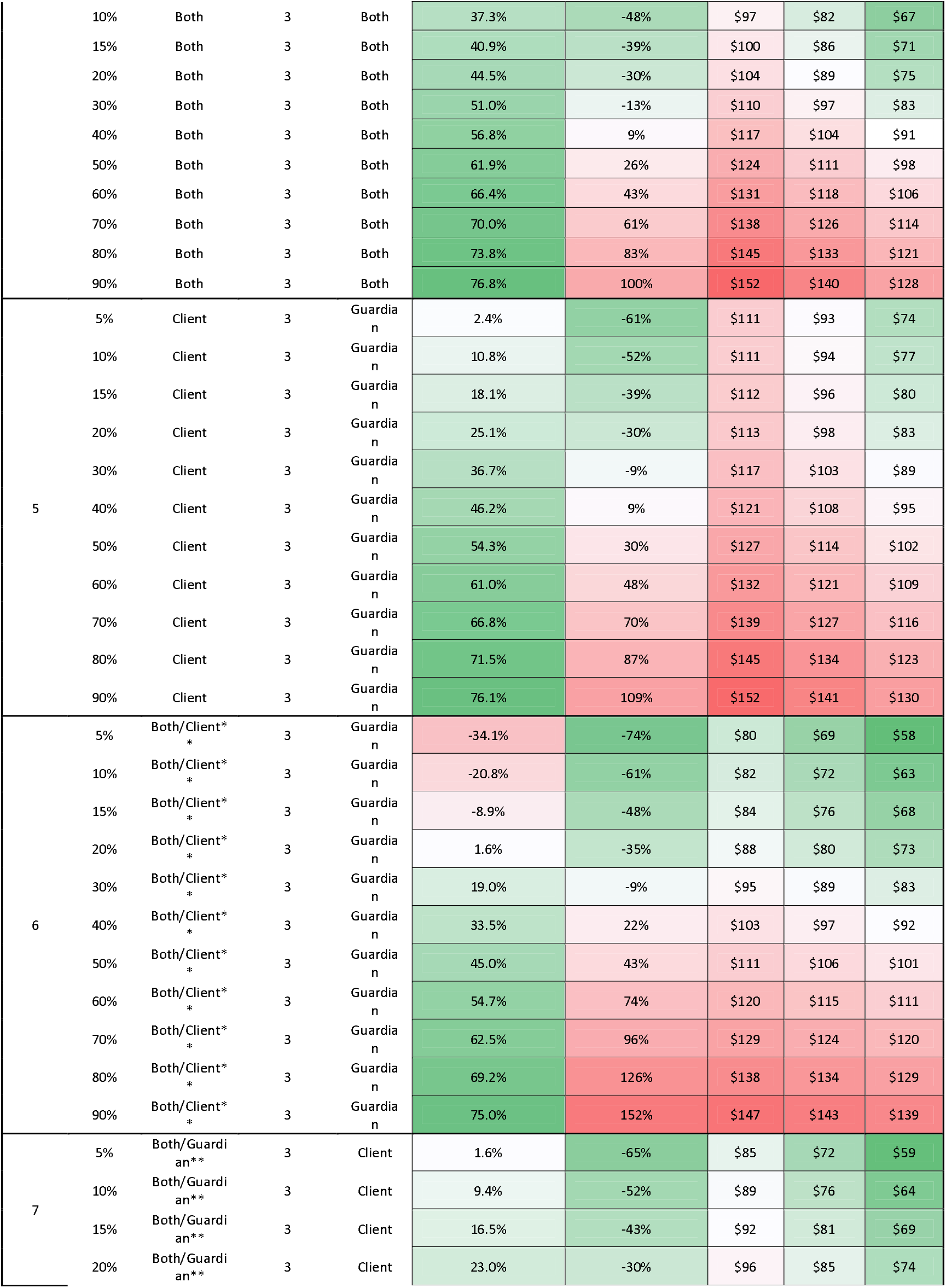
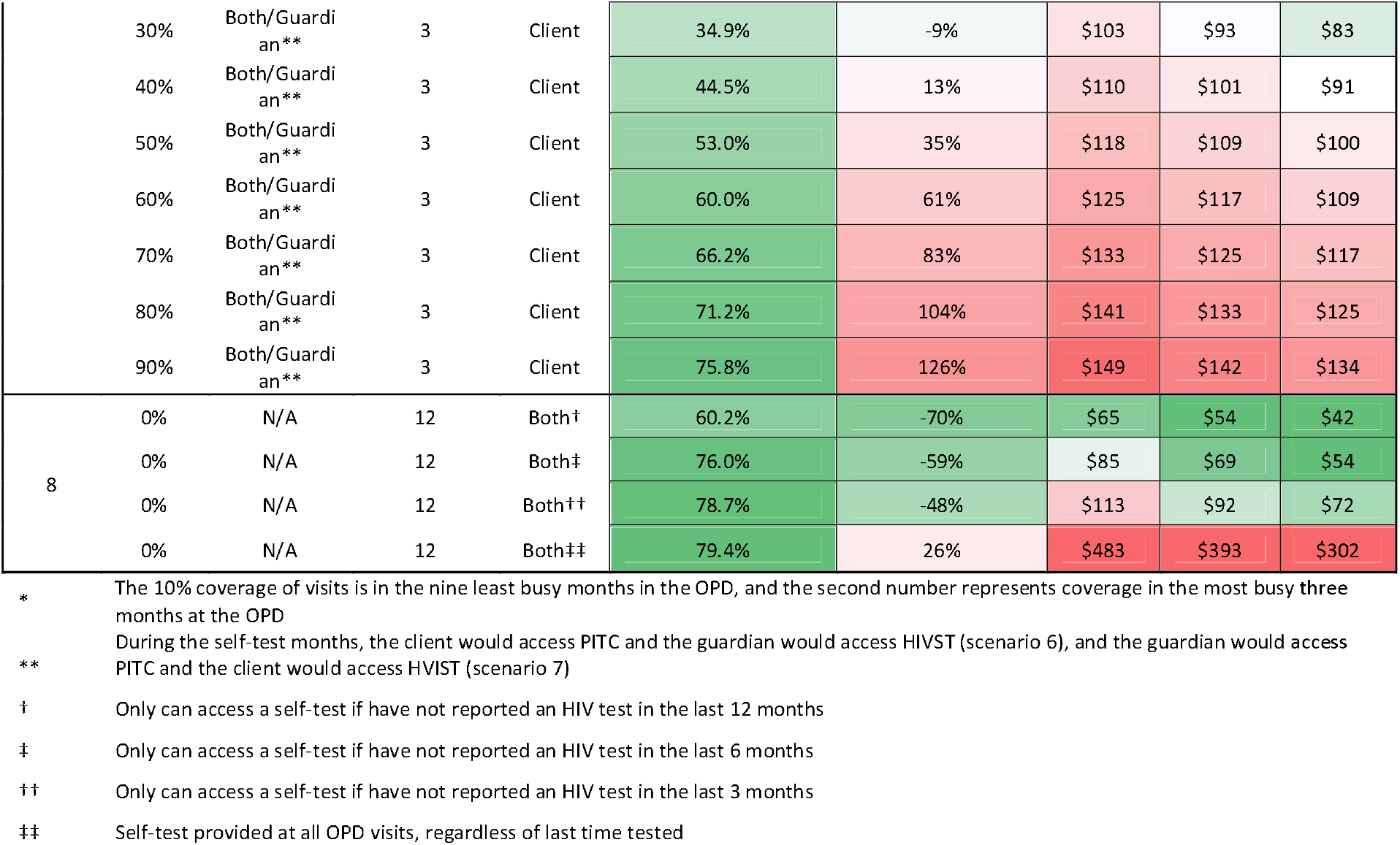
Proportion of men and youth tested and cost per new positive identified, by testing scenario, at different price points of the HIV self-test kit.

Among the HIVST-only scenarios, limiting testing to only those who have not been tested in the past 12 months (Ministry of Health guidelines) would reduce testing coverage to 74% of individuals getting tested within a 12 month window (versus 83% with no restrictions). The reduction is largely due to when individuals’ visit health facilities and not returning after they would become eligible for testing (i.e., >12 months since last test). Limiting testing to those who have not been tested in the past 6 months increased the proportion tested to 81% of individuals getting tested within a 12 month window. There was limited marginal gain when further loosening testing eligibility criteria (82% and 83% of people tested when limiting testing to those who have not tested in the past 3 months and with no time restrictions on past testing, respectively).

At the highest HIVST kit price ($1.80), the only cost-saving scenario was where PITC covers 10% of all OPD client visits at random (for clients and guardians) and increases to 60% of all visits receiving PITC at random during the highest-volume three months of the year (Scenario 3, Table 4). The subsequent four scenarios on the cost-effectiveness frontier were all variations of Scenario 8 (i.e., 12 months of HIVST offered at the health facility): year-round HIVST offered, limited to those who have not tested in the last 12 months ($39/additional positive identified), limited to those who have not tested in the last 6 months ($262/additional positive identified), the last 3 months ($1,983/additional positive identified), and not limited by time since last test ($84,592/additional positive identified).

**Table 4.** Testing scenarios that are on the cost-effectiveness frontier at different levels of HIVST kit price

| Scenario | Percent of visits covered with PITC | Person targeted PITC | HIVST Number of months implemented | Person targeted HIVST | Total cost | Total positives identified* | Incremental cost-effectiveness ratio |
| --- | --- | --- | --- | --- | --- | --- | --- |
| Price of HIVST kit= \$1.80 | | | | | | | |
| 3 | 10%/60% | Both | - | - | \$167,741 | 2,095 | Cost-saving |
| 1 | 50% | Client | - | - | \$172,407 | 2,011 | Baseline** |
| 8 | 0% | N/A | 12 | Both† | \$213,407 | 3,269 | \$39 |
| 8 | 0% | N/A | 12 | Both‡ | \$303,550 | 3,591 | \$262 |
| 8 | 0% | N/A | 12 | Both†† | \$411,443 | 3,646 | \$1983 |
| 8 | 0% | N/A | 12 | Both‡‡ | \$1,769,456 | 3,662 | \$84,592 |
| Price of HIVST kit= \$1.40 | | | | | | | |
| 7 | 5% | Both/Guardian | 3 | Client | \$149,131 | 2,073 | Cost-saving |
| 1 | 50% | Client | - | - | \$172,407 | 2,011 | Baseline** |
| 8 | 0% | N/A | 12 | Both† | \$175,794 | 3,269 | \$22 |
| 8 | 0% | N/A | 12 | Both‡ | \$249,355 | 3,591 | \$228 |
| 8 | 0% | N/A | 12 | Both†† | \$337,161 | 3,646 | \$1,614 |
| 8 | 0% | N/A | 12 | Both‡‡ | \$1,438,165 | 3,662 | \$68,583 |
| Price of HIVST kit= \$1.00 | | | | | | | |
| 8 | 0% | N/A | 12 | Both† | \$138,181 | 3,269 | Cost-saving |
| 1 | 50% | Client | - | - | \$172,407 | 2,011 | Baseline** |
| 8 | 0% | N/A | 12 | Both‡ | \$195,160 | 3,591 | \$177 |
| 8 | 0% | N/A | 12 | Both†† | \$262,878 | 3,646 | \$1,245 |
| 8 | 0% | N/A | 12 | Both‡‡ | \$1,106,874 | 3,662 | \$52,573 |

\* Assuming 2.5% positivity
\*\* Not on cost-effectiveness frontier
† Only can access a self-test if have not reported an HIV test in the last 12 months
‡ Only can access a self-test if have not reported an HIV test in the last 6 months
†† Only can access a self-test if have not reported an HIV test in the last 3 months
‡‡ Self-test provided at all OPD visits, regardless of last time tested

At the second highest HIVST kit price ($1.40), the scenario in which PITC covers 5% of all visits at random during the year for both clients and guardians and provides HIVST to clients only during the highest-volume three months of the year for clients only (not guardians) (Scenario 7) is considered cost-saving compared to baseline PITC. Similarly to when the HIVST kit price was $1.80, the subsequent four scenarios on the cost-effectiveness frontier were all sub-scenarios of Scenario 8 (i.e., 12 months of HIVST offered at the health facility): limited to those who have not tested in the last 12 months ($22/additional positive identified), the last 6 months ($228/additional positive identified), the last 3 months ($1,614/additional positive identified), not limited by time since last test ($68,583/additional positive identified).

At the lowest HIVST kit price ($1.00), the scenario in which HIVST is offered for 12 months (limited to those who have not tested for HIV in the previous 12 months, Scenario 8) was considered cost-saving compared to baseline PITC, resulting in a 20% reduction in total costs compared to the current coverage of PITC alone. The subsequent three scenarios on the cost-effectiveness frontier were the remaining versions of Scenario 8: limited to those who have not tested in the last 6 months ($177/additional positive identified), last 3 months ($1,245/additional positive identified), not limited ($52,573/additional positive identified).

## DISCUSSION

In this data-driven, individual-based model, we assessed the cost-effectiveness of multiple implementation strategies at different HIVST price points for HIV testing in OPD settings in Malawi that would increase testing coverage among men and youth. We found that facility-based HIVST may increase community-level HIV testing coverage among men and youth when implemented in OPD settings. Men and youth are historically underserved by current PITC programs, as standard PITC achieves poor coverage within busy OPD settings.^3,11^ Implementing year-round HIVST in OPD settings was highly cost-effective in most scenarios- and cost-saving compared to the baseline PITC when the price of the HIVST kit is reduced to $1.00. The overall increase in community-level testing coverage is possible due to the high frequency in which men and youth visit health facilities either as a clients or guardians, but have not typically been offered testing services.

The only PITC-based scenarios that can increase the proportion of men and youth tested to the same degree as year-round HIVST require a significant increase in the number of personnel available. To that end, the reduction in health care worker time required for HIVST compared to the current status quo is a significant advantage to the HIVST scenarios, requiring up to 70% fewer personnel than the current standard of care—a critical advantage to the health system as human resources are scarce. Saved human resource capacity from HIVST implementation could be used in other activities such as patient linkage, treatment support, or provide support elsewhere in the facility.

While we may expect cost per new positive to increase the closer we get to achieving 95-95-95 due to the increased difficulty in reaching those not yet in care, careful implementation modelling and planning can assist in identifying how to maximize the use of limited financial and human capital resources.^2^ Our findings contribute to a growing body of evidence on how facility-based HIVST can contribute to achieving the UNAIDS first 95 goals for those traditionally unreached by HIV services. HIVST is consistently shown to increase testing rates in comparison with standard PITC models, even in randomised controlled trials where providers receive additional support for implementing PITC.^5,6,8,12^ There is consensus that HIVST is associated with a higher absolute number of patients on treatment for HIV, although linkage rates differ based on how HIVST is implemented,^5,8,12^ and ART initiation has been lower among those diagnosed through self-testing in comparison to PITC.^8^ Hybrid facility-based PITC + HIVST strategies may allow health facilities to maximize the benefits of both modalities, increasing coverage and reducing personnel costs without sacrificing quality of and linkage to care.

There are several limitations to our study. First, for different standard PITC coverage levels, we randomly assigned individuals in the model to be offered an HIV test, and the uptake of that test was dependent on their survey response. In reality, PITC coverage among outpatient visits is unlikely to be completely random. Any selection in favour of someone at increased risk of HIV infection would result in increased cost-effectiveness of any scenarios including PITC through a reduction in the cost per positive test. However, the sensitivity of any type of screening needs to be weighed against the additional time it takes to administer the screening, which could increase human resource requirements and offset the reduction in cost per positive test. Second, the model was based on self-reported survey data. Participants in the survey may have under or overestimated the number of visits to the health facility. This may impact the total estimated proportion of men and youth that would receive an HIV test, but not differentially by scenario. Third, we have assumed a uniform distribution of positive cases across all scenarios to calculate a cost per positive identified to ensure comparability to the literature. While it is possible that some scenarios are more/less likely to identify new positives with fewer tests, given that the target population is in every instance identical (men and youth in the OPD) the yield is unlikely to differ meaningfully. Finally, due to people reporting health facility visits retrospectively, it is likely that dates of more recent visits are more precise as compared to visits that were further in the past. To circumvent this, we truncated our results to the past 12 months. Additionally, instead of modelling specific months, we modelled low- and high-volume OPD months to simulate coverage in either type of scenario.

To conclude, facility-based HIVST in the OPD is cost-effective and can significantly increase access to HIV testing for men and youth. The feasibility of covering all OPD visits with HIVST will depend on the available budget for test kits. Additional investment in capacity to implement year-round facility-based HIVST, limiting testing based on time-since-last-HIV-test, and introduction of lower-priced HIVST products should be prioritized to maximize the impact of facility-based testing strategies.

## Data Availability

Underlying model input data are available upon request.

